# Atypical action updating in a dynamic environment associated with adolescent obsessive-compulsive disorder

**DOI:** 10.1101/2021.11.08.21266056

**Authors:** Aleya A. Marzuki, Matilde M. Vaghi, Anna Conway-Morris, Muzaffer Kaser, Akeem Sule, Annemieke Apergis-Schoute, Barbara J. Sahakian, Trevor W. Robbins

## Abstract

**Background:** Computational research had determined that adults with obsessive-compulsive disorder (OCD) display heightened action updating in response to noise in the environment and neglect meta-cognitive information (such as confidence) when making decisions. These features are proposed to underlie patients’ compulsions despite knowledge they are irrational. Nonetheless, it is unclear whether this extends to adolescents with OCD as research in this population is lacking. Thus, this study aimed to investigate the interplay between action and confidence in adolescents with OCD.

**Methods:** Twenty-seven adolescents with OCD and 46 controls completed a predictive-inference task, designed to probe how subjects’ actions and confidence ratings fluctuate in response to unexpected outcomes. We investigated how subjects update actions in response to prediction errors (indexing mismatches between expectations and outcomes) and used parameters from a Bayesian model to predict how confidence and action evolve over time. Confidence-action association strength was assessed using a regression model. We also investigated the effects of serotonergic medication.

**Results:** Adolescents with OCD showed significantly increased learning rates, particularly following small prediction errors. Results were driven primarily by unmedicated patients. Confidence ratings appeared equivalent between groups, although model-based analysis revealed that patients’ confidence was less affected by prediction errors compared to controls. Patients and controls did not differ in the extent to which they updated actions and confidence in tandem.

**Conclusions:** Adolescents with OCD showed enhanced action adjustments, especially in the face of small prediction errors, consistent with previous research establishing ‘just-right’ compulsions, enhanced error-related negativity, and greater decision-uncertainty in paediatric-OCD. These tendencies were ameliorated in patients receiving serotonergic medication, emphasising the importance of early intervention in preventing disorder-related cognitive deficits. Confidence ratings were equivalent between young patients and controls, mirroring findings in adults OCD research.

## Introduction

Intrusions and compulsions displayed by individuals with obsessive-compulsive disorder (OCD) are ego-dystonic in nature (Sasson et al., 1997), as they occur despite being at odds with the core beliefs of the sufferer. Patients repeatedly engage in maladaptive washing and checking behaviour, for instance, despite having awareness that they are excessive and irrational.

Congruently, adults with OCD do not rely on meta-cognitive information, such as confidence, to inform decisions (Vaghi et al., 2017). Additionally, adult patients continue to respond to devalued stimuli despite being aware that the stimuli are no longer predictive of outcomes (Apergis-Schoute et al., 2017; Gillan et al., 2014; Vaghi et al., 2019). One explanation for this disconnect between actions and beliefs is that patients experience elevated uncertainty surrounding how events result from specific actions (Fradkin, Adams, Parr, Roiser, & Hupperts, 2020). They mistrust or place less weight on prior evidence in their decision-making and hence carry out compulsive behaviours that are at odds with pre-existing information to cope with the uncertainty. Supporting this theory, adults with obsessive-compulsive traits are found to rely less on past feedback information when making decisions on a probabilistic learning task, which led them to regard otherwise expected outcomes as ‘surprising’ (Fradkin, Ludwig, Eldar & Huppert, 2020).

Importantly, Vaghi and colleagues (2017) used a predictive-inference task to formally probe the interplay between meta-cognition (confidence) and action when environments are uncertain. Adults with OCD were tasked with predicting where a coin would land on a screen and rating how confident they were in their predictions. The task was probabilistic in that the coin predominantly landed in the same location with occasional deviations. Patients appropriately updated their confidence ratings based on changes in the coin location, but their actions did not reflect this knowledge. Instead, actions were driven by the most recent observation (where the coin landed most recently) instead of accumulated information (where the coin landed most frequently). Authors concluded that those with OCD can develop an accurate internal model of the task environment but fail to use this knowledge to guide their actions. In addition, computational modelling of the patients’ data revealed that correct confidence updating was uncoupled from excessive actions, further confirming that meta-cognition does not influence their actions.

No research has assessed whether beliefs and actions are updated abnormally in adolescents with OCD. Nonetheless, evidence has emerged that paediatric patients show enhanced ‘decision thresholds’ during certain tasks, wherein they continue to sample information prior to making a choice even when sufficient information has already been acquired (Erhan et al., 2017; Hauser et al., 2017b). This is suggestive of a dissociation between action and knowledge, as young patients continuously seek information even when doing so no longer has value.

Curiously, youths with OCD do not show cognitive impairments to the same extent as their adult counterparts (Abramovitch et al., 2015; Marzuki et al., 2020). As a result, devising a cognitive model of OCD that can account for both child and adult subtypes is challenging. This is in part due to the limited research conducted on youth-OCD samples compared to adult samples. As atypical meta-cognition is a consistent finding in adult-OCD research, it is important to understand whether this is also the case in adolescents with OCD. If so, it could serve as a potential shared impairment underpinning OCD symptoms in both childhood and adulthood.

Thus, this study aimed to investigate the relationship between confidence and action in a sample of adolescents with OCD using the predictive-inference task originally devised by McGuire, Nassar, Gold, & Kable (2014) and employed by Vaghi et al. (2017) to test adults with OCD. Based on prior findings in adult-OCD, we hypothesised, that compared to healthy age-matched controls, adolescent patients’ decisions would be driven by most recent feedback instead of information accumulated across the task, despite appropriate confidence ratings. We also predicted that adolescents with OCD would reveal an action-confidence dissociation on the task.

## Methods

### Sample

Seventy-three youths (12-19 years) completed the Predictive-Inference task (46 controls, 27 patients). Table 1 summarises the demographic and clinical characteristics. Eleven patients from the OCD group were receiving SSRIs (selective serotonin reuptake inhibitors) at the time of the study while 16 were unmedicated. Eight patients were receiving sertraline (Mean [M] = 118.75; standard deviation [SD] = 53.03) and 3 were receiving fluoxetine (M = 36.67; SD= 15.28). Groups were matched for gender, age, and IQ (intelligence quotient). However, the OCD group had significantly elevated depression, anxiety, and obsessive-compulsive severity scores compared to CTLs (Table 1).

**Table 1:** Mean scores and standard deviations per group and measure.

|  | **CTL(n = 46)** | **OCD(n = 27)** | **Statistic** |
| --- | --- | --- | --- |
| GENDER(F:M) | 28/18 | 18/9 | $\chi^2(1)=0.25, p = 0.62$ |
| AGE | 16.59(1.78) | 16.16(1.67) | $Z = 1.81, p = .07$ |
| WASI-II (IQ)<sup>a</sup> | 107.61(11.62) | 107.12(13.02) | $t(70) = 0.17, p = .87$ |
| BDI** | 46.46(5.27) | 59.03(9.55) | $t(35.45) = -6.31, p < .001$ |
| BAI** | 45.98(7.66) | 65.48(9.37) | $Z = -6.88, p < .001$ |
| OCI** | 8.13(6.49) | 31.70(13.92) | $Z = -6.35, p < .001$ |
| CY-BOCS <sup>a</sup> | N/A | 23.62(4.84) | N/A |
| CY-BOCS<br>Obsessions <sup>a</sup> | N/A | 11.23(2.18) | N/A |
| CY-BOCS<br>Compulsions <sup>a</sup> | N/A | 12.35(2.99) | N/A |

Details of questionnaires in Supporting Information. Mean(SD) reported. Key- CTL: Control Group; OCD: Obsessive-Compulsive Disorder group; WASI-II: Wechsler's Abbreviated Scale of Intelligence – II; IQ: Intelligence Quotient; BDI: Beck's Depression Inventory(t-scored); BAI: Beck's Anxiety Inventory(t-scored); OCI: Obsessive-Compulsive Inventory; CY-BOCS: Child Yale-Brown Obsessive-Compulsive Scale. N/A: Not applicable. \* $p < .05$ ; \*\* $p < .01$ ; <sup>a</sup>missing data from one OCD participant.

### Ethical considerations

This study was approved by the East of England - Essex Research Ethics Committee (REC 10/H030149/49). All volunteers gave written informed consent before beginning testing and were compensated at the rate of £8 per hour. Parental consent was also obtained if participants were under 16 years old.

### Inclusion and Exclusion Criteria (FigS1)

Adolescents with OCD were recruited via Child and Adolescent Mental Health Services around the United Kingdom. Healthy controls were recruited via advertisements in state secondary schools and on noticeboards around Cambridgeshire. Patients were screened by an experienced psychiatrist to rule out comorbid psychiatric and neurological conditions in a clinical interview supplemented by the Mini International Neuropsychiatric Interview (MINI, Sheehan et al., 1998; Sheehan et al., 2010).

To qualify for the study, those in the OCD group had to meet DSM-5-TR (Diagnostic and Statistical Manual of Mental Disorders-5-Text Revision) diagnostic criteria for OCD, have OCD as their primary diagnosis, and score 12 or above on the Children’s Yale-Brown Obsessive-Compulsive Scale 3. Apart from OCD, other significant Axis I psychiatric disorders were exclusion criteria. Those with severe physical impairments affecting eyesight or motor performance were also excluded, as they were predicted to affect task performance. Control participants (CTLs) were also screened to ensure they had no history of neurological or psychiatric illness.

### Predictive-Inference Task

Participants were instructed to make predictions about where ‘coins’ emitting from the centre of a circular ring would land by positioning an orange ‘bucket’ on the same circular ring to catch them (Figure 1). After positioning the bucket, participants rated their confidence in their choice on a scale (1-100). Before the task began, participants were informed that coins mostly flew to approximately the same location, but that location could change sometimes. No time limit was imposed but participants were instructed to respond as quickly as possible.

**Figure 1:**
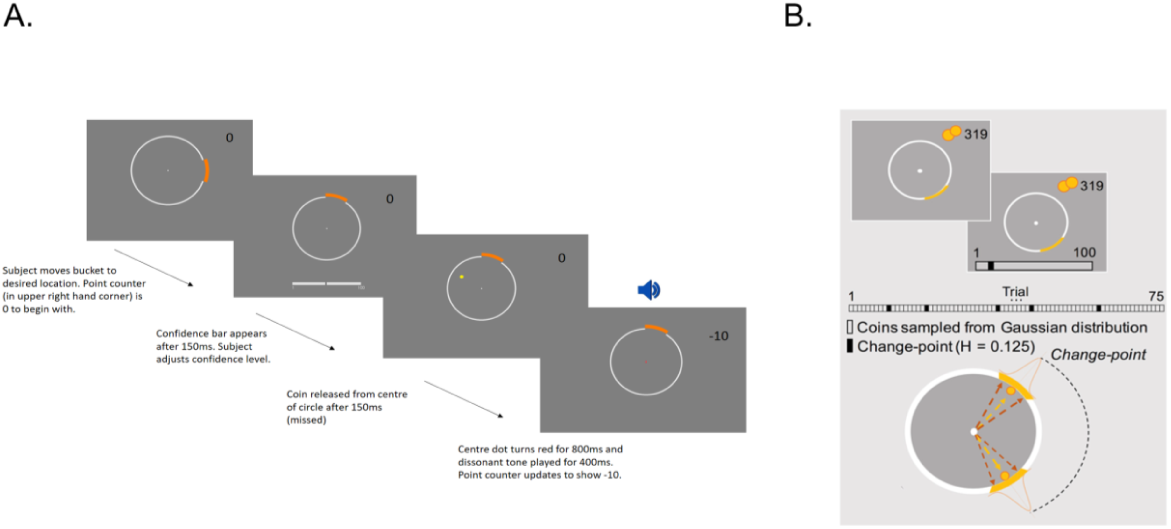
Predictive-Inference Task. A. An example of a trial where subject predicted coin location incorrectly. B: Coin locations are determined using a Gaussian distribution on most trials. When a change point occurs, the coin location changes drastically according to a uniform distribution. The probability of a change point occurring at any point during the task was 0.125. Black bars in the figure above represent change point trials. Otherwise, coins are sampled from a Gaussian distribution (white bars). Figure B adapted with permission from Vaghi et al. (2017).

The location the coin would be released to in each trial was mostly determined by sampling a Gaussian distribution. Hence, coins landed in a similar location with small variations driven by noise. The mean of this distribution usually remained stable over a block of trials but changed at random intervals (change-points, Figure 2) when it was resampled from a uniform distribution. The probability of a change-point occurring at any point in a block was set to 0.125. Thus, participants were required to form a new belief about the mean of the Gaussian distribution each time a change-point occurred. Conversely, they had to maintain the same belief about the mean of the Gaussian distribution when small changes in coin position were due to noise. There were 360 possible locations for coins to fly to when a change point occurred.

**Figure 2:**
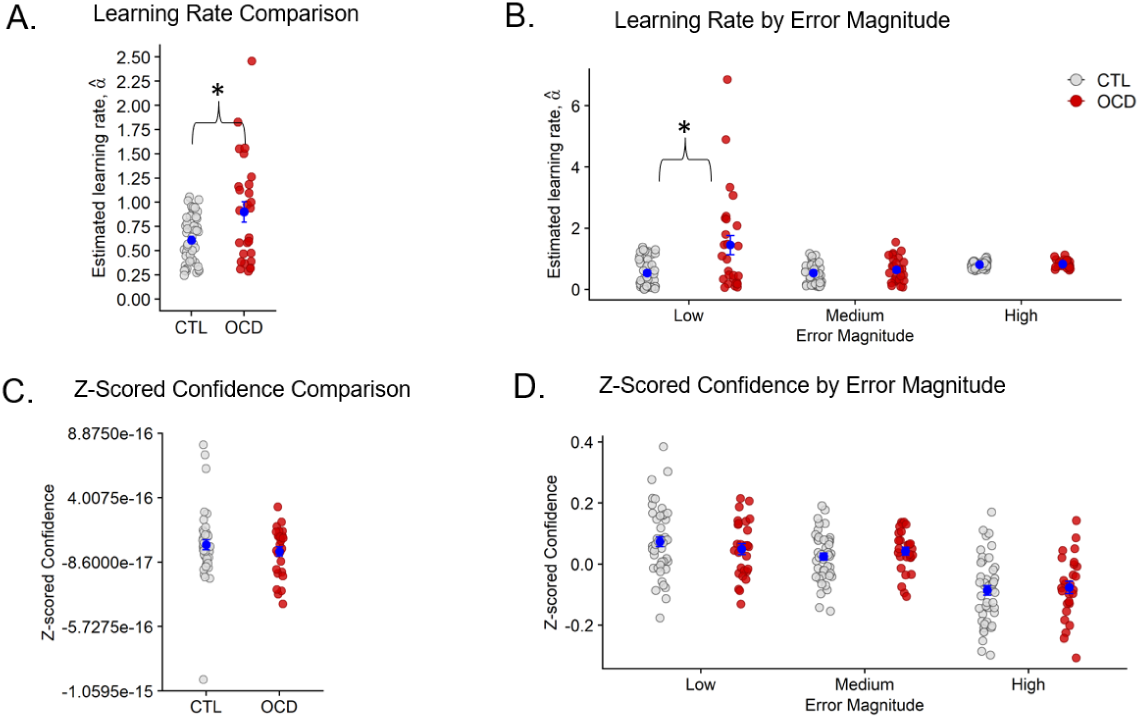
Learning rate and confidence. Note: *p < .05. A: The OCD group showed significantly higher learning rates compared to CTLs. B: Learning rates were significantly enhanced in the OCD group only when prediction errors were low. C and D: There were no significant group differences for mean Z-scored confidence and when confidence was binned by prediction error.

During the task, the bucket could be moved around the ring using a Griffin PowerMate USB rotary controller. Participants confirmed responses by pressing the spacebar on the laptop keyboard. After 150 milliseconds (ms), participants would be prompted to rate their confidence. A coin was then released for 150ms. If the coin landed within the boundaries of the bucket, participants were awarded 10 points. If the coin landed outside the boundaries, they lost 10 points. Stimuli were presented to participants using Matlab R2017b and Psychtoolbox v.3. Participants completed one practice block of 20 trials and 4 blocks of 75 trials each in the main task. The task lasted approximately 20 minutes.

### Statistical Analysis

Learning Rate and Confidence Analysis

Data manipulation and statistical analysis were conducted in Matlab R2017b and RStudio 3.5.0. Analysis and statistical models were adapted from Vaghi et al.’s (2017).

For each participant, learning rates on every trial (α_t_) were computed to understand how evidence accumulated in the task’s noisy environment influenced participants’ actions (positioning of the bucket):

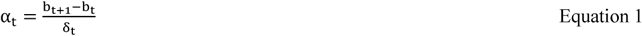

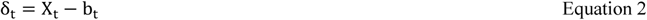

In Equation 1 and 2, b_t_ and b_t+1_ are the chosen bucket positions at trial t and trial t+1 respectively. δ_t,_, the prediction error, is the difference between the location of the particle (X_t_) at trial t and the position of the bucket at trial t (b_t_). Trials where the estimated learning rate (α) exceeded the 95^th^ percentile (calculated separately for each group) or where prediction error = 0 were excluded from the analysis. This type of filtering was employed as extreme values are reported to be due to noisy processes other than error-driven learning (Nassar et al., 2010). As several trials in our sample revealed extremely high learning rates (α_t_>1), our exclusion threshold was more stringent than what was employed by Vaghi et al. (2017) who only excluded trials exceeding the 99^th^ group percentile.

A two-sample t-test was used to confirm that there was no significant difference in the proportion of trials removed between the OCD and CTL groups (Supporting Information – Overall Data Checks). Difference in mean learning rates between groups was analysed using a Wilcoxon rank-sum test.

Raw confidence ratings were converted to z-scores and a two-sample t-test was used to assess group differences.

Next, we assessed whether magnitude of prediction error (the difference between expectations and outcomes, Equation 2) had a significant effect on learning rates and confidence. The learning rates and z-scored confidence ratings were divided into 3 quantiles based on magnitude of prediction error (low, medium, and high) using the ‘quantile’ function in Matlab. For each prediction error quantile, the mean learning rate and z-scored confidence were computed separately per group (OCD vs CTL).

Due to homogeneity of variance and normality violations, the Welch-James test from the ‘welchADF’ package in RStudio (Villacorta, 2017) was used to determine the effects of group on learning rates, depending on the magnitude of prediction error (low, medium, high). Post-hoc paired Wilcoxon tests with Bonferroni correction were conducted following the main Welch-James test.

The effects of group and prediction error on confidence were assessed using a Mixed-ANOVA. The Hyun-Feldt correction was applied due to violations of the sphericity assumption.

### Influence of Model Parameters on Learning and Confidence

We used a quasi-optimal Bayesian learning model to simulate data. The same model was also fitted to participant data (Supporting Information for model parameters and formulations). We compared participant’s actions and confidence to that of the Bayesian model by inspecting the average learning rates and confidence ratings 4 trials before and 4 trials after a change point occurred.

Linear regressions were conducted to estimate how much participants updated their actions and confidence over time according to parameters from the Bayesian model. The parameters included absolute prediction error (|δ_t_| absolute difference between belief [where bucket was positioned] and location of coin at each trial), change point probability (the relative likelihood that the coin is sampled from a new distribution, i.e., that a change-point has occurred), and relative uncertainty (reflects uncertainty regarding the mean of the coin distribution). These parameters were inserted as predictor variables in the regression models. Hit/Miss was also included as a categorical variable in the regressions to assess whether action and confidence were influenced by immediate feedback.

To assess the effects of Bayesian model parameters on participants’ actions, the dependent variable, ‘action’ was calculated by multiplying the learning rate, α_t_ by the absolute prediction error, |δ_t_| The predictors in this model were also multiplied by |δ_t_| Within the confidence regression model, z-scored reported confidence (not multiplied by |δ_t_|) was included as the dependent variable. Predictors associated with *t – 1* were inserted in the model, as confidence would be affected by information associated with the immediate previous trial. The last trial of each block per participant was removed before conducting the regressions as learning rates could not be estimated from these trials. Goodness-of-fit of each model to participant data was assessed by calculating median R-squared values.

To formally examine whether confidence and action are dissociated in adolescent patients, a third linear regression model was conducted with absolute confidence update (absolute difference between z-scored confidence scores on trial t and t-1) as the independent variable and absolute action update (absolute difference between where bucket was positioned at trial t and t-1) as the dependent variable. If confidence and action were linked, increased adjusting of the bucket position would correspond to a similar magnitude of change in confidence ratings.

To ascertain the presence of group differences in performance, beta coefficients associated with each predictor across the three regressions were extracted and compared between patients and controls. Independent sample t-tests were employed for these comparison analyses. Wherever the homogeneity of variance assumption was violated the Welch’s independent t-test was used instead. If the normality assumption was violated the Wilcoxon rank-sum test was used.

### Effect sizes

For t-tests, Cohen’s *d* was calculated as a measure of effect size whenever a significant effect was detected (small: <= 0.2 - <0.5, moderate: 0.5 > - < 0.8 and large: >= 0.8) (Cohen, 1988).

For Wilcoxon rank-sum tests, effect size was determined via the Wilcoxon effect size (Wilcoxon r) calculated by dividing the test z-statistic by the square root of the sample size (Z/√N). The following interpretations of effect size values were used, small: 0.1 - < 0.3, moderate: 0.3 - < 0.5 and large: >= 0.5 (Tomczak & Tomczak, 2014).

### Medication Analysis

The analyses described above were repeated subdividing the OCD group into unmedicated patients (MED-), and patients medicated with SSRIs (MED+). These exploratory analyses were conducted to investigate whether patients’ action and confidence were modulated by SSRIs.

## Results

### CTL vs OCD Analysis

The OCD group (M = 0.90; SD = 0.55) showed increased learning rates (Figure 2A) compared to CTLs (M = 0.61; SD = 0.25), Z = -2.18; *p* = .029, Wilcoxon’s r = 0.26. Next, our analysis of the effects of prediction error on learning rates revealed a significant main effect of prediction error magnitude T_wj_(2,34.87) = 73.73, *p* < .001. Learning rates were greater at high (M = 0.81; SD = 0.11) compared to medium (M = 0.57, SD = 0.34) prediction errors (Z =-5.61; *p =* 1.99e-08; Wilcoxon’s r = 0.44).

Importantly, a significant group-by-prediction error interaction was detected (Figure 2B), T_wj_(2,34.87) = 5.78, *p* = .0068. Post-hoc Wilcoxon tests revealed that OCD (1.45 ± 1.62) displayed enhanced learning rates compared to CTLs (0.53 ± 0.46) in response to low prediction errors (Z = -2.79; *p* = .0055; Wilcoxon’s r = 0.32). There were no group differences in learning rates at medium (CTL: M = 0.53, SD = 0.30; OCD: M = 0.64; SD = 0.40; *p* = .34) and high (CTL: M = 0.80, SD = 0.11; OCD: M = 0.82, SD = 0.13; *p* = .77) prediction errors.

In contrast, Z-scored confidence ratings (Figure 2C) did not differ significantly between groups (CTL: M = 4.50e-17; SD = 2.58e-16; OCD: M = -6.07e-18, SD = 1.85e-16; Z = 0.42, *p* = .68). When analysing effects of prediction error on confidence, we found a significant main effect of prediction error (F[2,142] = 32.11, *p* < .001). Participants predictably, reduced their confidence when prediction errors were high (M = -0.08, SD = 0.11) but increased in confidence when prediction errors were medium (M = 0.03, SD = 0.07) and low (M = 0.06, SD = 0.10), (high vs medium: t[72] = 5.97, *p* < .001; high vs low: t[72] = 6.61, *p* < .001; low vs medium: *p* > .05). There was no significant effect of group (F[1,71] = 0.05, *p* = .83) nor a significant group-by-prediction error interaction (F[2,142] = 32.11; *p* = .52) –Figure 2D.

### Regression Models

The median r-squared values for the action regression model were CTL: 0.87 and OCD: 0.80, while the median r-squared values for the confidence regression model were considerably lower (CTL: 0.085, OCD: 0.0643) indicating poor model fit.

There were no group differences in any of the beta values corresponding to parameters in the action model (pairwise comparisons: *p* > .05). In the confidence model, CTLs’ confidence was more influenced by prediction errors (Beta Coefficients: M = -0.086, SD = 0.13) compared to that of OCD participants (Beta Coefficients: M = -0.013, SD = 0.16), t[71] = -2.12; *p* = .037; Cohen’s *d* = 0.51. There were no other group differences in parameter values in the confidence model (pairwise comparisons: *p* > .05). The action and confidence regression results are summarised in Table S1 and S2 respectively.

Finally, the confidence-action regression model revealed no group differences in degree of action-confidence coupling (CTL: M = 0.053, SD = 0.064; OCD: M = 0.044, SD = 0.079; t[71] = 0.54, *p* = .59).

As learning rates were abnormally increased in the OCD group specifically during small prediction errors, we also applied the regression analyses to low prediction error magnitude trials only. However, no significant group differences were detected for model parameters (Tables S3 and S4).

### Analysis of Medication Effects

There were no significant differences in gender, age, and IQ between CTL, MED-, and MED+ groups (Table S5).

We excluded trials with learning rates that were greater than the 95^th^ percentile for each of the 3 groups, identical to what was done for the OCD vs CTL analysis. A Kruskal-Wallis test on learning rates showed a significant group effect, χ2(2) = 8.44, *p =* .015 (Figure 3A). MED- (M = 0.97, SD = 0.48) displayed higher learning rates than CTL (M = 0.61, SD = 0.25), Z= -3.09; *p*=.007; Wilcoxon’s r = 0.38, but not MED+ (M = 0.79, SD = 0.65), (MED+ = MED-, *p*=.69; MED+ = CTL, *p*=1.00). Z-scored confidence ratings were comparable across the 3 groups (CTL: M = 4.11e-17, SD = 2.57e-16; MED-: M = 3.39e-17, SD = 3.21e-16; MED+: M = - 4.25e-17, SD = 3.28e-16; χ2[2] = 4.34, *p* = .11).

**Figure 3:**
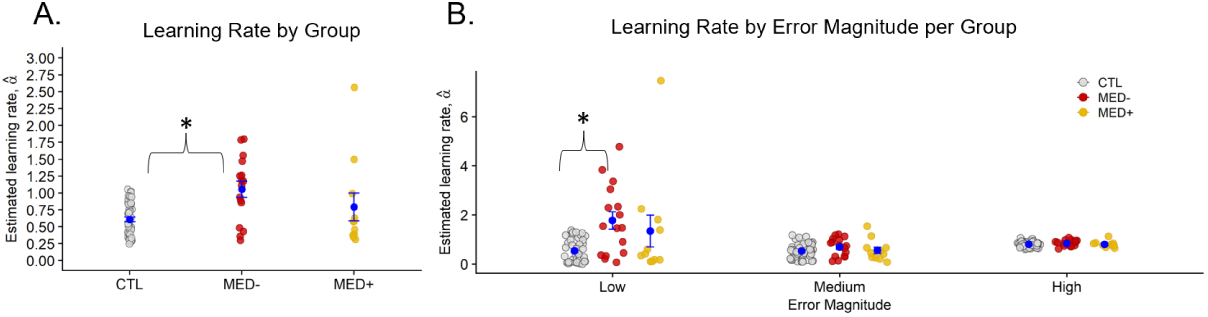
Learning rate by medication. A: The MED- group showed significantly increased learning rates compared to CTLs. There were no significant differences between MED- and MED+, and MED+ and CTLs. B: MED- showed significantly higher learning rates following low prediction errors compared to CTLs, but not MED+ patients. No group differences observed when considering medium and high prediction error trials.

After dividing learning rates by prediction error, we found a significant effect of group (T_wj_[2,17.22]=5.12, *p* =.018), prediction error (T_wj_[2,20.06]=66.23, *p* < .001, and a significant group-by-prediction error interaction (T_wj_[4,19.81]=4.22, *p* = .012). MED- (M = 1.78, SD = 1.42) showed higher learning rates at low prediction errors compared to CTL (M = 0.53, SD = 0.46), Z = -3.69, *p* < .001, Wilcoxon’s r = 0.45. Learning rates at low prediction errors were equivalent between MED+ (M = 1.35, SD = 2.16) and CTLs (*p* = .70), as well as between MED- and MED+ (*p* = .40) –Figure 4B.

**Figure 4:**
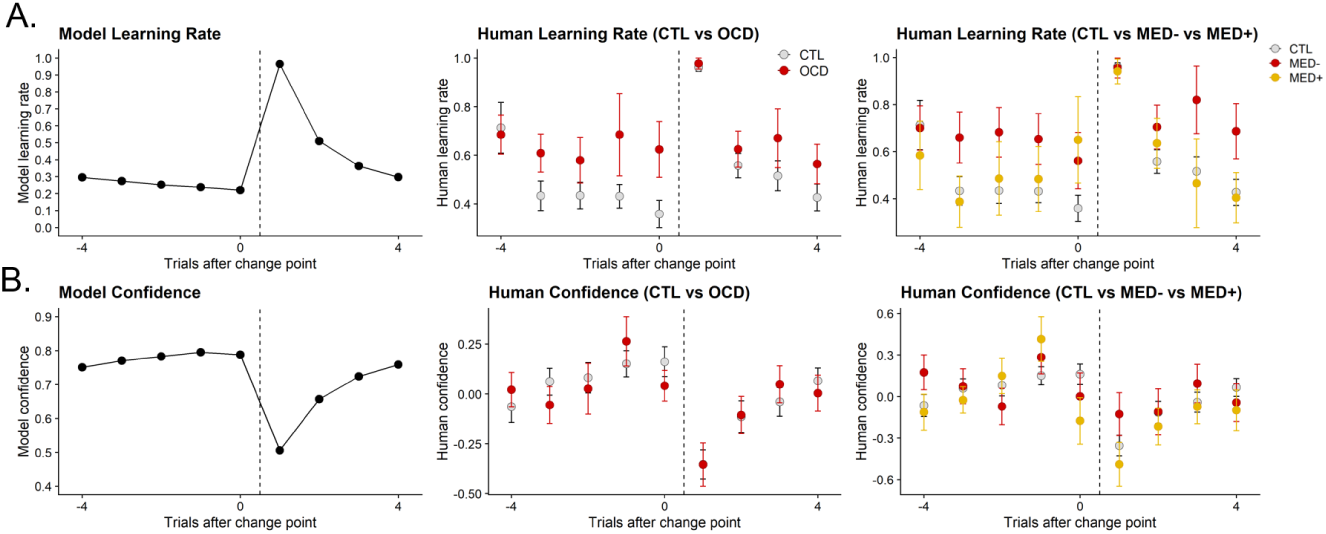
Comparing behaviour between the quasi-optimal Bayesian model and human participants following change points. A: Both Bayesian model and participants displayed increased learning rates following change points. However, the OCD group revealed elevated learning rates prior to change points compared to CTLs. The MED- group also showed higher learning rates than CTLs on trials preceding and following change points. B: The Bayesian model and human participants both decreased confidence ratings following change points. There was no observable difference between groups.

Action and confidence models revealed no significant group effects (Tables S6 and S7).

### Comparing model and human behaviour

Learning rate and confidence trajectories displayed by the quasi-optimal Bayesian model matched those observed in human participants. After change points occurred (signalling the need to discard old beliefs in favour of new information), participants suitably reacted by increasing their learning rates and decreasing their confidence ratings, hence demonstrating similar behaviour to that of the Bayesian model. However, the OCD group revealed higher learning rates than CTLs before a change point occurred. MED- also showed increased learning rates before and after change points occurred compared to CTLs. Confidence patterns were equivalent between all participant groups.

Correlations between task measures and clinical/intelligence scores were quantified using Pearson’s correlations (Table S8).

## Discussion

This study investigated the relationship between action and confidence in adolescents with OCD compared to healthy adolescents. The paradigm used was previously implemented by Vaghi et al., (2017) who uncovered increased action updating following recent feedback and a novel dissociation between action and confidence in adults with OCD. As predicted, adolescent patients in our study displayed increased action updating following recent feedback, most prominently when prediction errors were low. This was driven primarily by unmedicated patients. By contrast, there was no difference in confidence updating between patients and controls, although, in our model-based analysis, the adolescent OCD group’s confidence ratings were significantly less influenced by prediction errors. Contrary to our prediction, adolescents with OCD did not display a significant dissociation between action and confidence.

### Increased learning rates in OCD

While prior findings in adults with OCD indicated increased learning rates regardless of prediction error magnitude (Vaghi et al., 2017), current results showed that adolescents with OCD primarily updated actions excessively following low prediction errors. Enhanced updating at low prediction errors might indicate that patients are ‘tracking’ the location of the coin by moving the bucket every time the coin makes a small deviation from its last location. Perhaps the adolescents with OCD are pre-occupied with ensuring the coin lands with high certainty in the bucket. This is evocative of not-just-right obsessions (where a sufferer feels that their environment is not as it should be) often associated with OCD. In fact, majority of paediatric OCD patients describe having not-just-right related obsessions (Nissen & Parner, 2018). Trials where the coin just barely lands in the bucket may have triggered a not-just-right perception in patients, leading to the urge to re-arrange the bucket. Indeed, ordering/symmetry-related compulsions are strongly associated with not-just-right perceptions (Coles et al., 2003).

Additionally, frequent choice corrections observed in adolescents with OCD are consistent with research reporting increased error-related negativity (ERN) in paediatric OCD patients (Marzuki et al., 2020 for review). Importantly, error signals are generated in the absence of feedback and are triggered by a person’s own awareness that an error has indeed occurred (Gehring, Goss, Coles, Meyer, & Donchin, 1993). Heightened ERN in OCD could be likened to an internal ‘alarm bell’ that frequently sounds regardless of the degree of volatility in the external environment. In line with this, Fradkin et al.’s (2020a) computational model of OCD proposes that feeling excessively uncertain about the otherwise stable environment leads to patients perceiving that their rituals are not performed adequately, culminating in a tendency to repeat actions.

Moreover, excessive updating could be a form of atypical information gathering, which has been observed prior in youths with OCD on information sampling (Hauser et al., 2017b) and perceptual decision-making tasks (Erhan et al., 2017). Increased perceptual uncertainty could be driving this need to gather more information. Alternatively, patients may be taking a longer time than controls to learn the exact optimal location to place the bucket, which would be consistent with research suggesting that youths with OCD have learning deficits (Gottwald et al., 2018).

### Confidence Results

Normal confidence updating suggests that adolescent patients can construct a relatively accurate internal model of the task, but do not appear to use this information to guide their actions, mirroring adults with OCD (Vaghi et al., 2017).

However, model-based analysis revealed that the OCD group’s confidence ratings were insensitive to the influence of prediction errors, while controls appropriately decreased their confidence ratings when predictions errors were high. We caution that these results should be interpreted carefully given that 1) the confidence model fit was sub-par and 2) our model-free results showed comparable confidence ratings between patients and controls.

Confidence updating that is insensitive to prediction errors supports the proposal by Fradkin et al. (2020a) that individuals with OCD do not use external evidence to inform their beliefs. The finding is also reminiscent of recent research revealing that healthy adults with compulsive traits and intrusive thoughts display inflated confidence (Rouault et al., 2018; Seow & Gillan, 2020). Although, Seow and Gillan (2020) reported that compulsive participants’ confidence ratings were not strongly influenced by feedback (hit/miss), change point probability, and relative uncertainty, while our model analysis showed a reduced effect of only prediction error on patients’ confidence.

### No group differences in confidence-action coupling

Despite observing elevated learning rates but relatively normal confidence ratings in adolescents with OCD, and previous studies reporting a mismatch between action and confidence in obsessive-compulsive adults (Hauser et al., 2017a; Rouault et al., 2018; Seow & Gillan, 2020; Vaghi et al., 2017; Vaghi et al., 2019) we found no group differences when formally testing the strength of association between action and confidence.

The results suggest adolescents with OCD indeed update action and confidence in tandem and that perhaps confidence and action only become dissociated with age. An alternative explanation is that meta-cognition could still be developing in healthy adolescents, resulting in a lack of noticeable group differences and the deficit only apparent during adulthood. This is supported by research demonstrating that accurate meta-cognition only emerges in early adolescence but strengthens over time (Fandakova et al., 2020; Moses-Payne, Habicht, Bowler, Steinbeis, & Hauser, 2020; Weil et al., 2013). There is also computational evidence showing healthy adolescents do not use confidence to inform their decision-making (Jepma, Schaaf, Visser, & Huizenga, 2020). Hence, perhaps detecting an effect of OCD in our study is difficult as the adolescent control group are also updating action and confidence independently.

### Medication effects

We found that excessive updating was driven by unmedicated patients with OCD, while patients medicated with SSRIs did not differ significantly from controls. Thus, importantly, SSRIs appear to be ameliorating compulsive tracking of the coin by adolescent patients on the task.

This is consistent with research suggesting that medicated adult patients show superior performance to medication-naïve patients on various learning and planning tasks (Lochner et al., 2020; Palminteri et al., 2012). Importantly, SSRIs administered to adolescents and children with OCD is associated with significant improvements in verbal memory, processing speed, inhibition, and cognitive flexibility (Andrés et al., 2008).

## Conclusions

Prompted by previous work detailing a novel action-outcome dissociation in adults with OCD, we demonstrate that adolescents with OCD deviated most from healthy adolescent behaviour when experiencing small prediction errors where they made frequent action updates. Computational modelling revealed patients’ confidence ratings were not as influenced by prediction errors as those of control participants. We posit that youths with OCD update their actions according to internal factors that need to be researched further, rather than following observable changes in the task environment. This is consistent with prior research reporting uncertainty-driven information sampling and error-related negativity in this clinical population. We also provide evidence for aberrant action-updating to be remediated by SSRI treatment in youths with OCD, emphasising the importance of early intervention in improving disorder-related decision-making deficits.

## Key points

- Adults with OCD are known to show a dissociation between action and confidence, consistent with OCD being an ego-dystonic disorder.
- We found, for the first time, that adolescents with OCD update their actions excessively when changes in the environment were small.
- Patients’ confidence was updated similarly to control subjects.
- There is possible positive effect of SSRI medication on action updating.
- Patients’ actions may be driven by internal uncertainty rather than following observable changes in the task, reminiscent of real-life obsessions and compulsions which are at odds with reality.

## Data Availability

All data produced in the present study are available upon reasonable request to the authors

