## Supplementary Information for "Atypical action updating in a dynamic environment associated with adolescent obsessive-compulsive disorder"

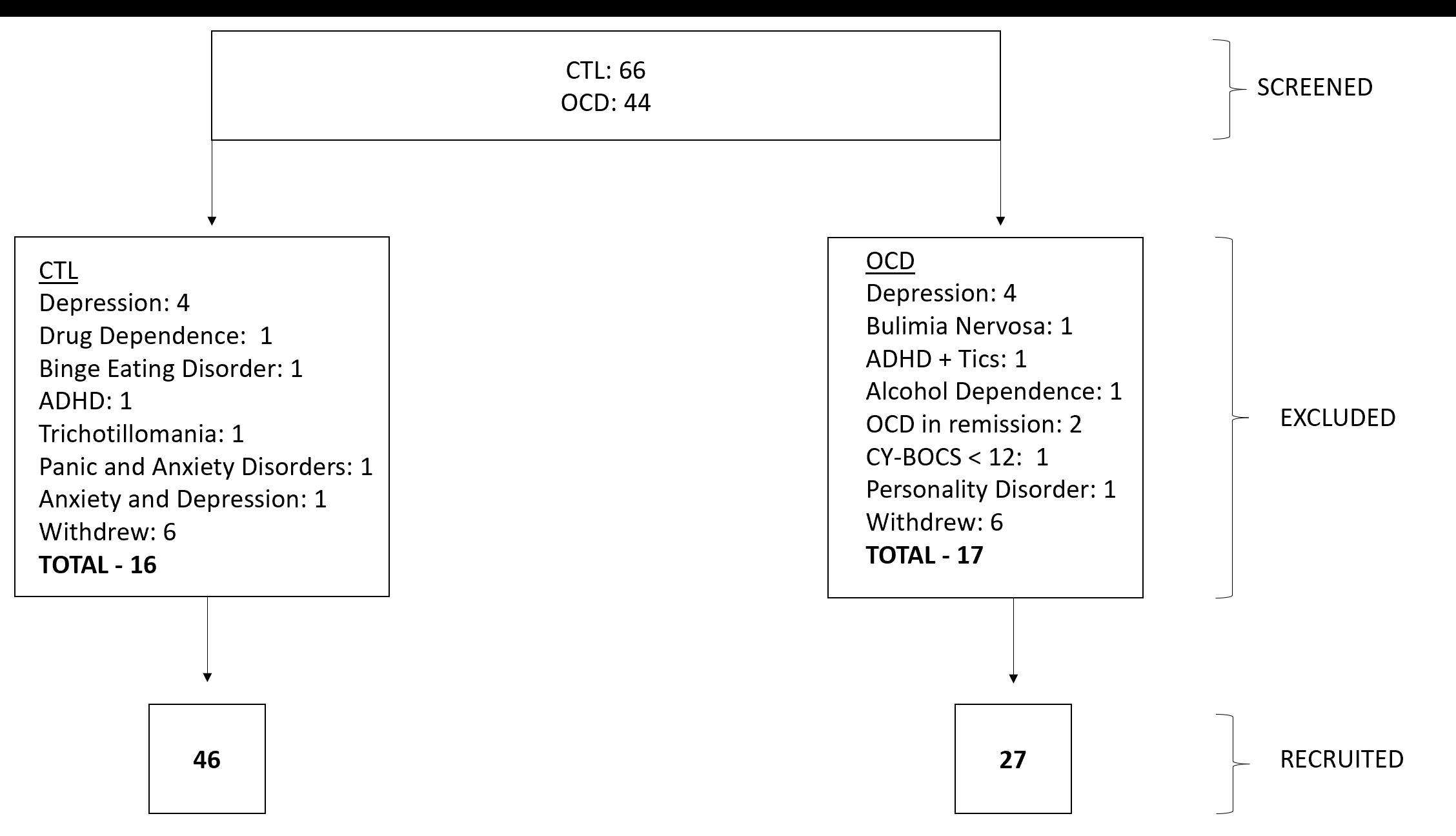

Figure S1: Screening and recruitment details for sample. Key- CTL: Control Group; OCD: Patient Group; ADHD: Attention Deficit Hyperactivity Disorder; CY-BOCS: Children’s Yale-Brown Obsessive-Compulsive Scale.

### Questionnaires

Self-reported obsessive-compulsive traits were assessed using the Obsessive-Compulsive Inventory-Revised [OCI-R (Foa et al., 1998)]. OCD symptom severity was also assessed with the Children’s Yale-Brown Obsessive-Compulsive Scale [CY-BOCS (Scahill et al., 1997)]. The CY-BOCS was only administered to participants with OCD. To obtain a measure of anxiety and depression, all participants completed the Beck Anxiety Inventory for Youth and the Beck Depression Inventory for Youth (Beck et al., 2005). IQ measures were obtained using the Wechsler’s Abbreviated Scale of Intelligence, Second Edition [WASI-II (Wechsler, 1999)]. The Full-Scale IQ-2 subtests (FSIQ-2) from the WASI-II were used comprising the Vocabulary and Matrix Reasoning tests.

### Computational Model

To better understand factors influencing trial-by-trial confidence and action updates in participants, a quasi-optimal Bayesian model (previously implemented by Vaghi et al., 2017) was fit to participant data. The model in question is termed a quasi-optimal Bayesian model as it attempts to approximate the behaviour of a full Bayesian learner which would infer future outcomes using information accumulated over all previous outcomes (Nassar et al., 2010), but using the less computationally complex delta rule. The delta rule (see Equation S1) involves updating a future belief (at t+1) based on the current belief and the error made in predicting the most recent outcome ($\delta_{t})$. The influence of the new outcome over beliefs (B_t_) is controlled by a learning rate ($\alpha_{t})$. At $\alpha_{t}=0$, the model’s belief is not changed following incoming outcome information at all, while at $\alpha_{t}=1$ the most recent outcome will completely influence the model’s beliefs.

$B_{t+1}=B_{t}+\alpha_{t} x \delta_{t}$	Equation S1

The prediction error, $\delta_{t}$ is defined as the difference between the current belief about the coin’s location, B_t,_ and the actual location of the coin, X_t_:

$\delta_{t}$ = X_t_ – B_t_	Equation S2

In turn, the model’s learning rate, α_t_, is defined as:

α_t_ = Ω_t_ + (1 - Ω_t_) (1 – *v*_t_)	Equation S3

α, in the above equation, was dynamic meaning it could change from trial-to-trial (as opposed to α parameters in other reinforcement learning studies which are kept constant). Ω_t_ in Equation S3 represents the change-point probability, indicating how likely the model thinks a change in location has occurred. α_t_ increases when the model assumes a change-point is likely to have taken place. This change point probability, Ω_t_, is constructed as the relative likelihood that a new location will be drawn from the same Gaussian distribution (N) centred around the current belief, B_t_ of the model, or alternatively from a uniform distribution over 360 possible locations:

$\Omega_{t}=\frac{{(X}_{t}|1,360)H}{{(X}_{t}\left| 1,360 \right)H+N \left( X_{t} \right|B_{t}, {\sigma^{2}}_{t})(1-H)}$	Equation S4

H in the equation above represents the hazard rate (the actual probability that the mean distribution has changed at any given trial). H was fixed at 0.125. $\Omega_{t}$will be close to 1 when the probability of the sample coming from a uniform distribution is higher than the probability of it being drawn from a Gaussian distribution (indicating a surprising outcome).

${\sigma^{2}}_{t}$ in Equation S4 is the estimated variance of the predictive distribution and is influenced by the variance of the generative Gaussian distribution ${\sigma^{2}}_{N}$ (noise in the location of the sample before a change point has occurred) and model confidence, *v*_t_:

${{\sigma^{2}}_{t}}={\sigma^{2}}_{N}+\frac{(1-v_{t}){\sigma^{2}}_{N}}{v_{t}}$	Equation S5

Lastly, model confidence, *v*, in Equation S5 is always computed for the subsequent trial, and considers uncertainty arising from imprecise estimation of the mean of the sample location:

$v_{t+1}=\frac{{\Omega_{t}{\sigma^{2}}_{N}+\left( 1- \Omega_{t} \right)\left( 1-v_{t} \right){\sigma^{2}}_{t}+ \Omega_{t}\left( 1- \Omega_{t} \right)({\sigma^{2}}_{t}}v_{t})^{2}}{{\Omega_{t}{\sigma^{2}}_{N}+\left( 1- \Omega_{t} \right)\left( 1-v_{t} \right){\sigma^{2}}_{t}+ \Omega_{t}\left( 1- \Omega_{t} \right)({\sigma^{2}}_{t}}v_{t})^{2}+{\sigma^{2}}_{N}}$	Equation S6

In the equation above there are 3 terms included in the numerator (from left to right): 1) reflects the variance when a change point is thought to have occurred, 2) represents variance when no change point is assumed to have occurred, and finally 3) reflects a rise in uncertainty when the model is unsure about whether a change point has occurred or not. The 3 terms from the numerator are repeated in the denominator with an added variance term reflecting uncertainty arising from noise in the Gaussian distribution. When an unexpected change in the task environment occurs, model confidence will decrease, and as a result, the learning rate, α_t_ in Equation S3 will increase. Relative uncertainty was inserted as (1-$\Omega$)*(1-$v$) in the regression models reported in the main manuscript which is consistent with the term in Equation S3.

Tables S1 and S2 summarise results obtained from the Action and Confidence regression models for OCD vs CTL groups.

**Regression Results for CTL vs OCD Groups (Low Error Magnitude Trials Only)**

We isolated trials where spatial prediction errors were low in magnitude and applied the regressions to the data from these trials. We did not include ‘Hit/Missed’ as a variable in the action and confidence regressions as participants always managed to obtain ‘hits’ (where the coin successfully lands in the bucket) during low error magnitude trials. No significant group differences were detected (see Tables S3 and S4).

There was also no group difference in action-confidence association strength when considering only low error magnitude trials (CTL: Mean [M] = 0.056, standard deviation [SD] = 0.11, OCD: M = 0.055, SD = 0.088; t(71) = 0.060, *p* = .95).

**Regression Results for CTL vs MED- vs MED+ groups**

Demographic and clinical measures per group are summarised in Table S5. There were significant group effects on BDI (χ2(2) = 30.029, p= 3.02e-07), BAI (χ2 (2)=47.222 , p = 5.00e-11), and OCI (χ2 (2)=40.391 , p = 1.70e-09) scores. Post-hoc Dunn’s tests revealed that compared to CTL, MED+ and MED- groups had elevated depression (MED- vs CTL: *p* = 4.48e-09., MED+ vs CTL: *p* = 1.37e-06), anxiety (MED- vs CTL: *p =*6.80e-08, MED+ vs CTL: *p* = 7.58e-06), and obsessive-compulsive scores (MED- vs CTL: *p* = 6.80e-08, MED+ vs CTL: *p* = 7.58e-06). There were no differences on these measures between MED- and MED+ groups (all *p* > .05). The median r-squared values for the regression models were as follows: action regression, MED+ = 0.77, MED- = 0.83; confidence regression, MED+ = 0.075, MED- = 0.069

There were no significant group differences in beta values across all parameters included in the action and confidence regression models (see Table S6 and S7).

There were no group differences in beta values for the action-confidence coupling regression (CTL: M = 0.053, SD = 0.064, MED-: M = 0.037, SD = 0.070, MED+: M = 0.050, SD = 0.061; F(2,70) = 0.372, *p* = .69), indicating that the association strength between action and confidence were equivalent between all groups.

### Overall Data Checks

After filtering trials that exceeded the 95^th^ percentile per group, we ascertained that there were no statistical differences in proportion of trials removed for the OCD and CTL groups, (mean proportion removed: CTL - M = 0.064, SD = 0.044; OCD – M = 0.064, SD = 0.050; F(1,71) = 0, *p* = .97), as well as for CTL, MED-, and MED+ groups (mean proportion removed: MED-: M = 0.077, SD = 0.064, MED+: M = 0.051, SD = 0.037; F(2,70) =0.94, *p* = .40). We also checked that accuracy (proportion of ‘hits) was equivalent between CTL and OCD groups (CTL: M = 0.61, SD = 0.062; OCD: M = 0.60, SD = 0.065; t(71) = 0.98, *p* = .33), and between CTLs and medication groups (MED-: M = 0.61, SD = 0.060; MED+: M = 0.61, SD = 0.057, F(2,70) = 0.81, *p* = .45)

**Correlations**

We conducted Pearson’s correlations between demographic/clinical measures and task variables (learning rates overall, low error magnitude learning rates, z-scored confidence, and action-confidence regression betas) to understand whether such variables are linked to clinical characteristics. These are summarised in the Table S8. There were no significant correlations when considering CTL, OCD, MED-, and MED+ groups separately.

Table S1: Summary of parameters for CTL and OCD obtained from the regression model on action

| **Parameter** | **Group** | **Mean Beta Coefficient** | **Standard Dev.** | **Statistics** |
| --- | --- | --- | --- | --- |
| PE | CTL | 0.43 | 0.30 | t(71) = 0.077; *p* =.94 |
|  | OCD | 0.43 | 0.31 |  |
| CPP | CTL | 0.47 | 0.29 | t(71) = 0.35; *p* =.73 |
|  | OCD | 0.44 | 0.27 |  |
| RU | CTL | 0.71 | 0.67 | Z = -0.080; *p* = .94^§^ |
|  | OCD | 0.88 | 1.07 |  |
| Hit/Missed | CTL | -0.74 | 0.24 | t(71) = .20 , *p* = .84 |
|  | OCD | -0.75 | 0.30 |  |

Key- PE: prediction error, CPP: Change Point Probability, RU: Relative Uncertainty. A two -sample t-test was used for statistical comparison unless indicated otherwise. ^§^ Wilcoxon’s Tes

Table S2: Summary of parameters for CTL and OCD obtained from the regression model on confidence

| **Parameter** | **Group** | **Mean Beta Coefficient** | **Standard Dev.** | **Statistics** |
| --- | --- | --- | --- | --- |
| PE* | CTL | -0.086 | 0.13 | t(71) = -2.12; *p* = .037; Cohen’s *d* = 0.51 |
|  | OCD | -0.013 | 0.16 |  |
| CPP | CTL | -0.12 | 0.21 | t(71) = 1.36; *p* = 0.18 |
|  | OCD | -0.19 | 0.26 |  |
| RU | CTL | -0.17 | 0.15 | t(71) = -0.22; *p* = .82 |
|  | OCD | -0.16 | 0.18 |  |
| Hit/Missed | CTL | 0.16 | 0.12 | t(71) = 1.82; *p* = .072 |
|  | OCD | 0.11 | 0.12 |  |

Key- PE: prediction error, CPP: Change Point Probability, RU: Relative Uncertainty. **p* < .05 A two -sample t-test was used for statistical comparison unless indicated otherwise.

Table S3: Summary of parameters for CTL and OCD obtained from Action regression model for low error magnitude trials only

| **Parameter** | **Group** | **Mean Beta Coefficient** | **Standard Dev.** | **Statistics** |
| --- | --- | --- | --- | --- |
| PE | CTL | -0.29 | 0.62 | Z =-0.81; *p* =.42 |
|  | OCD | -0.16 | 0.53 |  |
| CPP | CTL | 1.09 | 0.53 | Z = 1.18; *p* =.24 |
|  | OCD | 0.94 | 0.44 |  |
| RU | CTL | 1.20 | 1.08 | Z =.30; *p* =.77 |
|  | OCD | 1.27 | 1.48 |  |

Key- PE: prediction error, CPP: Change Point Probability, RU: Relative Uncertainty. The Wilcoxon’s Rank Sum test was used for all group comparisons

Table S4: Summary of parameters for CTL and OCD obtained from Confidence regression model for low error magnitude trials only

| **Parameter** | **Group** | **Mean Beta Coefficient** | **Standard Dev.** | **Statistics** |
| --- | --- | --- | --- | --- |
| PE | CTL | -0.026 | 0.25 | t(71) = -1.92; *p* =.058 |
|  | OCD | 0.093 | 0.27 |  |
| CPP | CTL | -0.0076 | 0.25 | t(71) = 1.54; *p* =.13 |
|  | OCD | -0.11 | 0.28 |  |
| RU | CTL | -0.11 | 0.22 | t(71) = 0.20; *p* = .84 |
|  | OCD | -0.12 | 0.19 |  |

Key- PE: prediction error, CPP: Change Point Probability, RU: Relative Uncertainty. A two -sample t-test was used for statistical comparison unless indicated otherwise.

Table S5: Mean scores and standard deviations per group and statistical test.

|  | **CTL (n = 46)** | **MED- (n = 16)** | **MED+ (n = 11)** | **STATISTIC** | **PAIRWISE COMPARISONS** |
| --- | --- | --- | --- | --- | --- |
| SEX(F:M) | 28/18 | 13/3 | 5/6 | χ2(2)=3.83 , p = 0.15 | - |
| AGE | 16.59 (1.78) | 16.28 (1.60) | 15.98 (1.84) | χ2(2)=3.67 , p = 0.16 | - |
| WASI-II (IQ)^a^ | 107.61 (11.62) | 107.27 (13.64) | 106.9 (14.96) | F(2,69) = 0.016, *p* =0.98 | - |
| BDI ** | 46.46 (5.27) | 59.06 (9.60) | 59.00 (9.94) | χ2(2) = 30.029, *p* < .001 | CTL < MED- & MED+  MED- = MED+ |
| BAI ** | 45.98 (7.66) | 66.13 (9.68) | 64.54 (9.28) | χ2 (2)=47.222 , *p* < .001 | CTL < MED- & MED+  MED- = MED+ |
| OCI ** | 8.13 (6.49) | 32.00 (13.64) | 29.55 (13.63) | χ2 (2)=40.391 , *p* < .001 | CTL < MED- & MED+  MED- = MED+ |
| CY-BOCS ^a^ | N/A | 24.47 (4.92) | 22.45 (4.70) | t(24) = 1.05, *p* = 0.30 | N/A |
| CY-BOCS Obsessions ^a^ | N/A | 11.33 (2.64) | 11.09 (2.55) | t(24) = 0.28, *p* = .79 | N/A |
| CY-BOCS Compulsions ^a^ | N/A | 13.07 (2.55) | 11.36 (3.38) | t(24) = 1.47, *p* = .16 | N/A |

Key*:* Mean (SD) reported. CTL: Control Group; MED-: Unmedicated patient group; MED+: Medicated patient group; WISC-IV: Wechsler’s Abbreviated Scale of Intelligence – II; IQ: Intelligence Quotient; BDI: Beck’s Depression Inventory (t-scored); BAI: Beck’s Anxiety Inventory (t-scored); OCI: Obsessive-Compulsive Inventory; CY-BOCS: Child Yale-Brown Obsessive-Compulsive Scale. **p*<.05; ***p*<.01; ^a^ missing data from one MED- participant.

Table S6: Summary of parameters for CTL MED- and MED+ obtained from Action regression model

| **Parameter** | **Group** | **Mean Beta** | **Standard Dev.** | **Statistics** |
| --- | --- | --- | --- | --- |
| PE | CTL | 0.43 | 0.30 | F(2,70) = 0.45, *p* = .64 |
|  | MED- | 0.50 | 0.35 |  |
|  | MED+ | 0.38 | 0.34 |  |
| CPP | CTL | 0.47 | 0.29 | F(2,70) = 0.96, *p* =.39 |
|  | MED- | 0.36 | 0.29 |  |
|  | MED+ | 0.50 | 0.31 |  |
| RU | CTL | 0.71 | 0.67 | χ2(2) = 2.22 ; *p* = .33^§^ |
|  | MED- | 0.70 | 1.18 |  |
|  | MED+ | 1.04 | 0.94 |  |
| Hit/Missed | CTL | -0.74 | 0.24 | F(2,70) = 0.34; *p* =0.71 |
|  | MED- | -0.78 | 0.26 |  |
|  | MED+ | -0.70 | 0.35 |  |

Key- PE: prediction error, CPP: Change Point Probability, RU: Relative Uncertainty. A One-Way ANOVA was used for statistical comparison unless indicated otherwise. ^§^ Kruskal-Wallis Test

Table S7: Summary of parameters for CTL, MED-, and MED+ obtained from Confidence regression model

| **Parameter** | **Group** | **Mean Beta** | **Standard Dev.** | **Statistics** |
| --- | --- | --- | --- | --- |
| PE | CTL | -0.086 | 0.13 | F(2,70) = 2.02 , *p* = .14 |
|  | MED- | -0.0048 | 0.15 |  |
|  | MED+ | -0.057 | 0.15 |  |
| CPP | CTL | -0.12 | 0.21 | F(2,70) =0.91, *p* = .41 |
|  | MED- | -0.17 | 0.27 |  |
|  | MED+ | -0.21 | 0.25 |  |
| RU | CTL | -0.17 | 0.15 | F(2,70) =0.45; *p* =.64 |
|  | MED- | -0.14 | 0.17 |  |
|  | MED+ | -0.20 | 0.20 |  |
| Hit/Missed | CTL | 0.16 | 0.12 | F(2,66) = 2.11; *p* = .13 |
|  | MED- | 0.084 | 0.12 |  |
|  | MED+ | 0.13 | 0.12 |  |

Key- PE: prediction error, CPP: Change Point Probability, RU: Relative Uncertainty. A One-Way ANOVA was used for statistical comparison unless indicated otherwise.

Table S8: Correlations between Demographic/Clinical Measures and Task Outcome Measures – All Subjects

|  | Age | IQ | Anxiety | Depression | OCI |
| --- | --- | --- | --- | --- | --- |
| LR | N.S. | N.S. | r = 0.24  *p* = .041 | N.S. | N.S. |
| Low Error Magnitude LR | N.S. | N.S. | r = 0.23  *p* = .049 | N.S. | N.S. |
| Confidence (z-scored) | N.S. | N.S. | N.S. | N.S. | N.S. |
| Action-Confidence Regression Betas | N.S. | r = 0.29  *p* = .013 | N.S. | N.S. | N.S. |

Key- LR: Learning rates; N.S: Non-significant
